# Modeling and Sensitivity Analysis of Coronavirus Disease (COVID-19) Outbreak Prediction

**DOI:** 10.1101/2020.11.18.20234419

**Authors:** Ahmad Sedaghat, Seyed Amir Abbas Oloomi, Mahdi Ashtian Malayer, Shahab S. Band, Amir Mosavi, Laszlo Nadai

## Abstract

Susceptible-infectious-recovered-deceased (SIRD) model is an essential model for outbreak prediction. This paper evaluates the performance of the SIRD model for the outbreak of COVID-19 in Kuwait, which initiated on 24 February 2020 by five patients in Kuwait. This paper investigates the sensitivity of the SIRD model for the development of COVID-19 in Kuwait based on the duration of the progressed days of data. For Kuwait, we have fitted the SIRD model to COVID-19 data for 20, 40, 60, 80, 100, and 116 days of data and assessed the sensitivity of the model with the number of days of data. The parameters of the SIRD model are obtained using an optimization algorithm (lsqcurvefit) in MATLAB. The total population of 50,000 is equally applied for all Kuwait time intervals. Results of the SIRD model indicate that after 40 days, the peak infectious day can be adequately predicted. Although error percentage from sensitivity analysis suggests that different exposed population sizes are not correctly predicted. SIRD type models are too simple to robustly capture all features of COVID-19, and more precise methods are needed to tackle the correct trends of a pandemic.

## I. Introduction

Since the outbreak of COVID-19 in Wuhan, China, in December 2019, 215 countries worldwide reported the pandemic cases of COVID-19 summing total 9,655,329 diagnosed cases, total 488,136 death cases, and total 5,244,462 recovered cases by 26 June 2020 [1]. Kuwait reported a total of 42,788 diagnosed, current 9,082 under treatment, 152 critical, 339 deceased, 33,367 recovered, and 23 quarantined on 26 June 2020 by the ministry of health (MOH) [2]. Kuwait government have already removed full lockdown except for some highly susceptible areas and taken steps to ease on most of the closures across the country. Accurate prediction of COVID-19 development is proved to be very difficult in many countries due to unstable trends of the pandemic in these countries. To show the uncertainty of SIRD prediction on the dynamics of COVID-19 [3], we have assessed an optimized SIRD model based on time intervals of COVID-19 developments.

In this paper, we have used SIRD model optimized by an optimization algorithm (lsqcurvefit) in MATLAB for fitting the model with COVID-19 data for several consequence time intervals in Kuwait. We have calculated the goodness of fit using the coefficient of determination (R2). The sensitivity of results on the size of COVID-19 days of data are presented and discussed for the pandemic outbreak in Kuwait, and conclusions of the work are drawn.

## II. Materials and methdos

SIR model is the simplest model used in epidemic/pandemic studies, which is expressed by a set of consequence ordinary differential equations (ODE) introduced by Kermack and McKendrick [4]. Details of 3 sets of ODEs can be found in numerous publications based on susceptible (S) cases, active infected cases (I), and removed cases (R) (including recovered and death) [5, 6].

### A. SIRD Model

We used SIRD model which includes one more equation on number of death (D) as follows [7]:

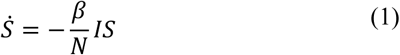

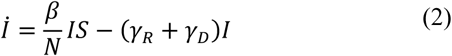

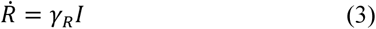

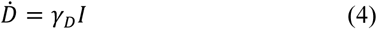

In equations (1-4), S is susceptible, I is the active infected, R is the recovered, and D is the death populations. Time derivatives are shown by over-dot; e.g.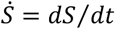. The transmission rate *β*, the recovery rate *γ*_*R*_, and the death rate *γ*_*D*_ are the unknown parameters of the SIRD model.

The removing rate is obtained by *γ* = *γ*_*R*_ + *γ*_*D*_. Reproduction number *R*_0_ = *β*/*γ* is an important characteristic parameter on dynamics of SIR model. Some studies suggest that if *R*_0_ < 1 then the nonlinear dynamics of the SIRD model is stable, and predictions are close to reality [8]. But, if *R*_0_ > 1, then the dynamical system of the SIRD model is unstable. In the SIRD model, total population N is considered as constant.

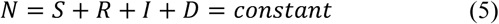

We should differentiate between total active infected (I) and total infected cases (IC) as follows:

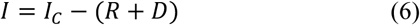

An example of initial conditions to solve the SIRD model is as follows:

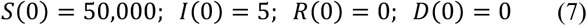

Optimized SIRD model parameters are obtained by fitting COVID-19 data using the curve-fitting optimization algorithm (lsqcurvefit) in MATLAB.

## III. Results

### A. Regression coefficient

Regression coefficient (R2) is applied for each of four populations, susceptible, infected, recovered, and death to check goodness of fitting SIRD model with COVID-19 data. The regression coefficient is a good measure on dealing with futurism prediction. The regression coefficient (R2) compares predicted values (y) against actual data (x) as follows [9]:

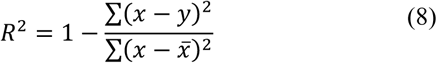

In equation (8),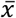 is the average of COVID-19 data values. The regression coefficient (R^2^) close to unity shows best fit.

### B. Sensitivity analysis

Having actual peak day values, SIRD model predictions using different time duration set data can be assessed to determine sensitivity of the model to size of COVID-19 data. A comparison of actual peak day values (*x*) and predicted values (*y*) are obtained in percentage of error as follows [10]:

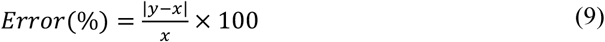

Sensitivity of SIRD model is obtained using equation (9) on COVID-19 days of data values. Closer error to zero percentage means less sensitivity of SIRD model to data size.

### C. SIRD Predictive Results for COVID-19 in Kuwait

Results of SIRD model using COVID-19 data are obtained for 20, 40, 60, 80, 100, and 116 days cases with optimized parameters from 24 February to 19 June 2020 as shown in Fig. Table 1 summarizes optimized parameters obtained corresponding to cases in Fig. 1. It is observed that 20 days data cannot provide any coefficient of regressions for different population due to lack of data. For instance, there was zero death cases during the first 20 days period so it is not possible to determine the regression coefficient. For 40- and 60-days interval, the death data was still insufficient to provide a meaningful accuracy.

**TABLE 1.** Optimized SIRD model parameters and accuracy for COVID-19 in Kuwait (19 June 2020).

| No. of days | Growth rate ($\beta$) | Recovery rate ($\gamma_R$) | Death rate ($\gamma_D$) | Removing rate ($\gamma$) | Re-production number ($R_0$) | $R^2(S)$ | $R^2(I)$ | $R^2(R)$ | $R^2(D)$ |
| --- | --- | --- | --- | --- | --- | --- | --- | --- | --- |
| 20 | 0.1831 | 0.01363 | 0.0044 | 0.01798 | 10.18 | - | - | - | - |
| 40 | 0.1451 | 0.03524 | 0.0022 | 0.03739 | 3.88 | 0.8141 | 0.6376 | 0.9581 | - |
| 60 | 0.1294 | 0.02308 | 0.0016 | 0.02464 | 5.25 | 0.9611 | 0.9396 | 0.9812 | - |
| 80 | 0.1396 | 0.04158 | 0.0014 | 0.04295 | 3.25 | 0.9915 | 0.9715 | 0.9824 | 0.5744 |
| 100 | 0.1340 | 0.03576 | 0.0015 | 0.0373 | 3.59 | 0.9970 | 0.9837 | 0.9817 | 0.7098 |
| 116 | 0.1519 | 0.05501 | 0.0011 | 0.056 | 2.71 | 0.9987 | 0.9117 | 0.9694 | 0.7841 |

**Figure 1.**
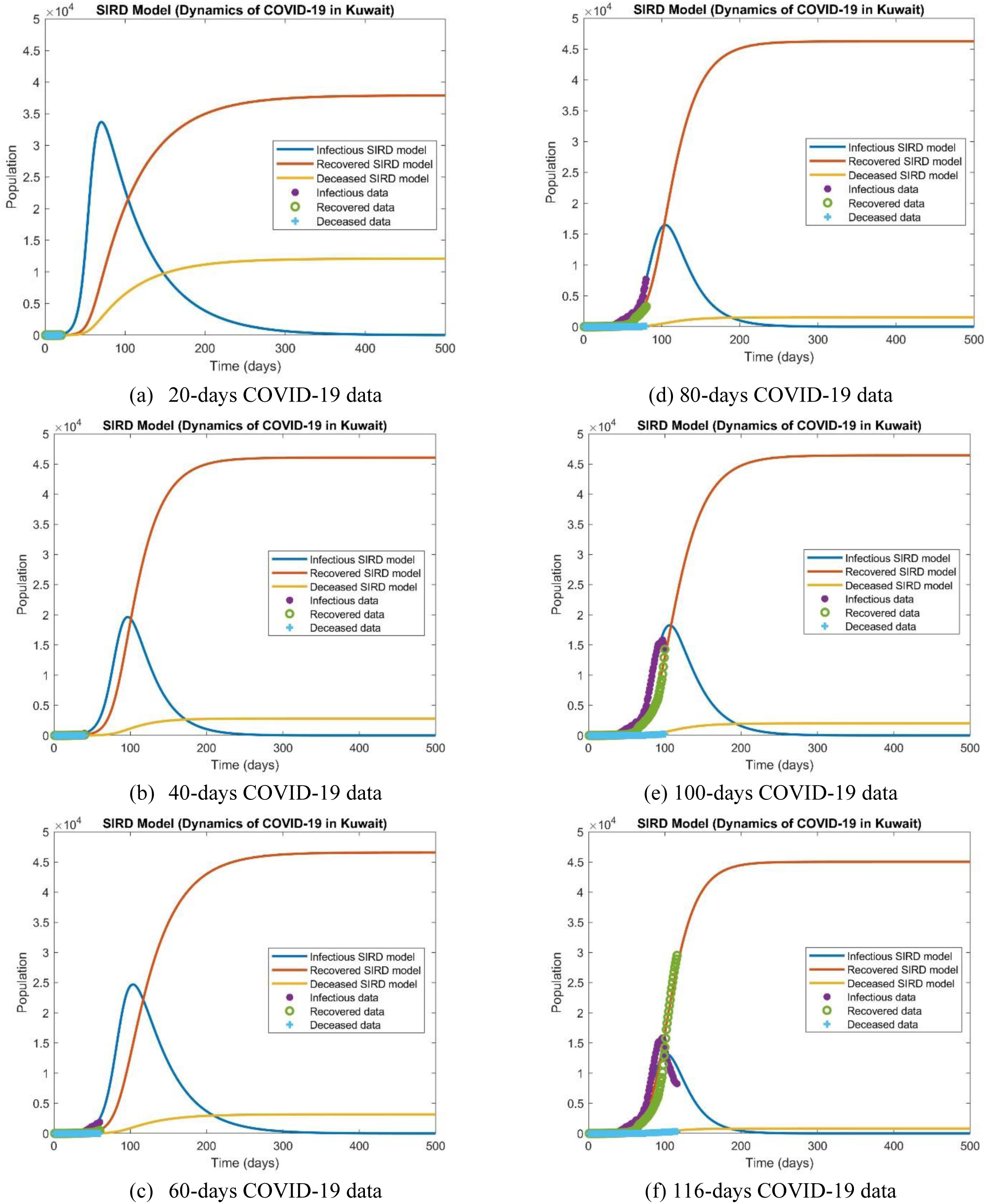
Optimized SIRD model predictions for dynamics of COVID-19 in Kuwait (19 June 2020).

Figure 2 compares values of susceptible, active infected, recovered, deceased, and total infected using various time duration of COVID-19 data for peak day of infection. As shown in Fig. 2, with 20, 40, 60, 80, 100, and 116 days cases of data, the peak infection dates 5/5, 1/6, 7/6, 9/6, 10/6, and 6/6/2020 are predicted, respectively. A linear trend is observed in all of these predictions for different populations; yet we do not get precise solution.

**Figure 2.**
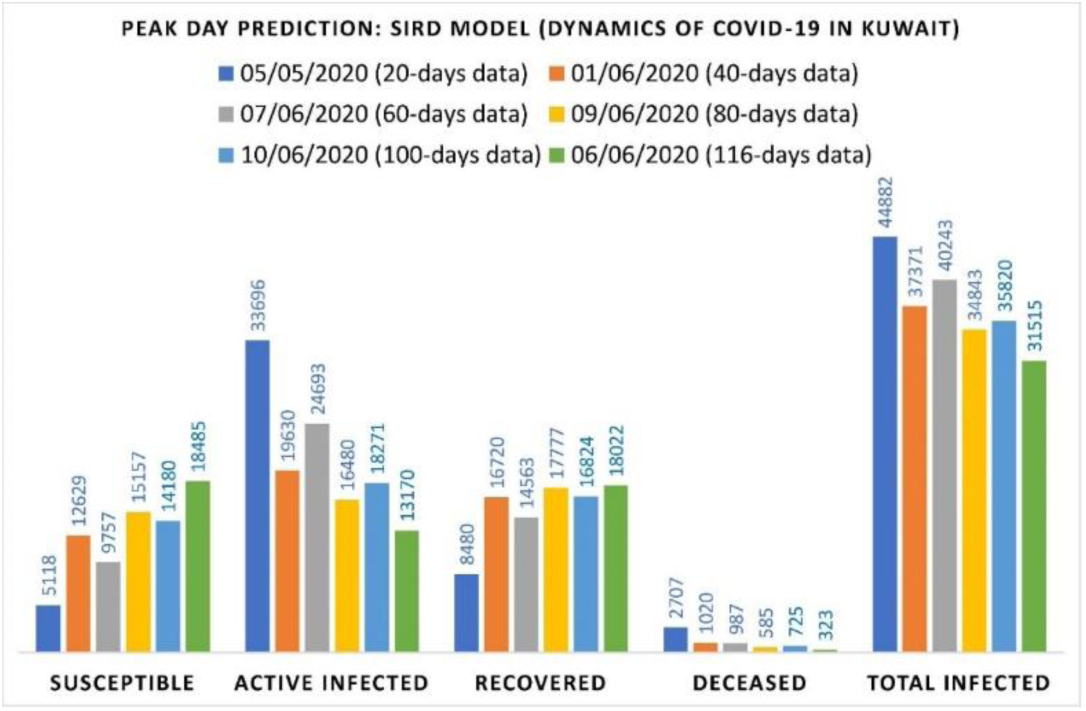
Peak infection day predictions using optimized SIRD model using 20, 40, 60, 80, 100, and 116 COVID-19 data cases (19 June 2020)

Figure 3 shows the trends of these solutions as COVID-19 days progressed. No convergence to certain limits is observed in this figure. Actual peak day of infection was observed on 30 May 2020 (96 days after 24 February 2020) according to worldometer [1].

**Figure 3.**
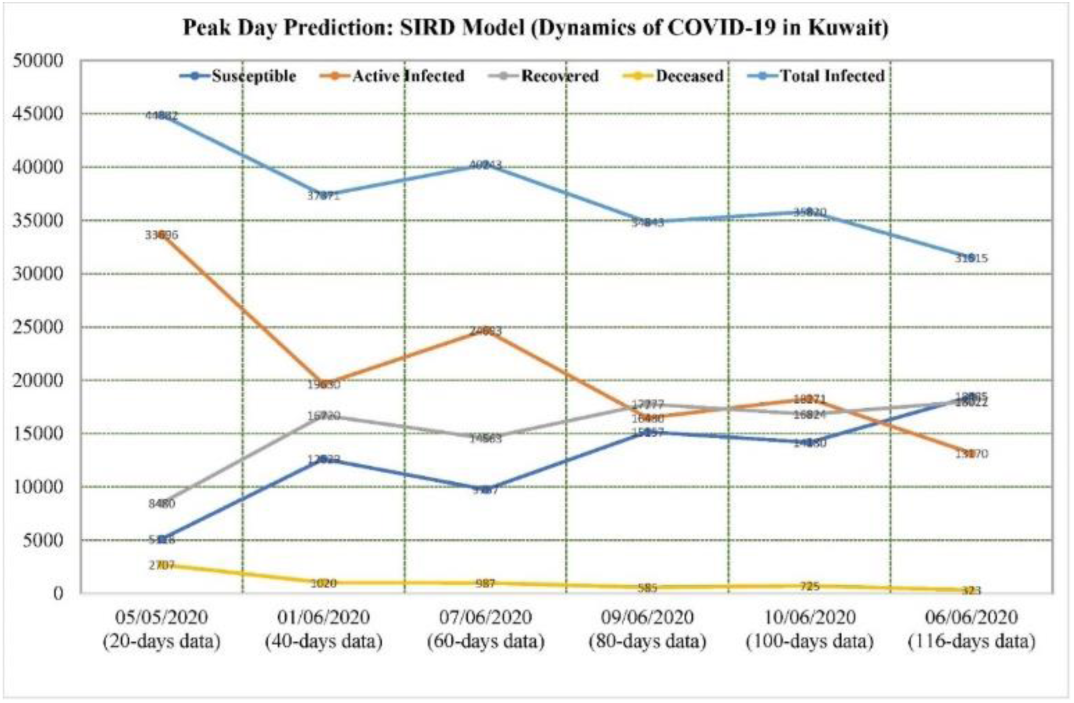
Population size predictions on peak infection day using optimized SIRD model for 20, 40, 60, 80, 100, and 116 COVID-19 data cases (19 June 2020).

The predicted and actual values of peak infectious day and corresponding errors using equation (6) are shown in Table 2. The lowest error is 20% on total infected cases, 57% on total deceased cases, and 17% on total recovered using 116 days of COVID-19 data cases. The results on Table 2 show poor predictive capability of SIRD model on precise prediction of size of populations in CVID-19 pandemic. But, SIRD model quickly and nearly accurately predicts the peak day of infection.

**TABLE 2.**
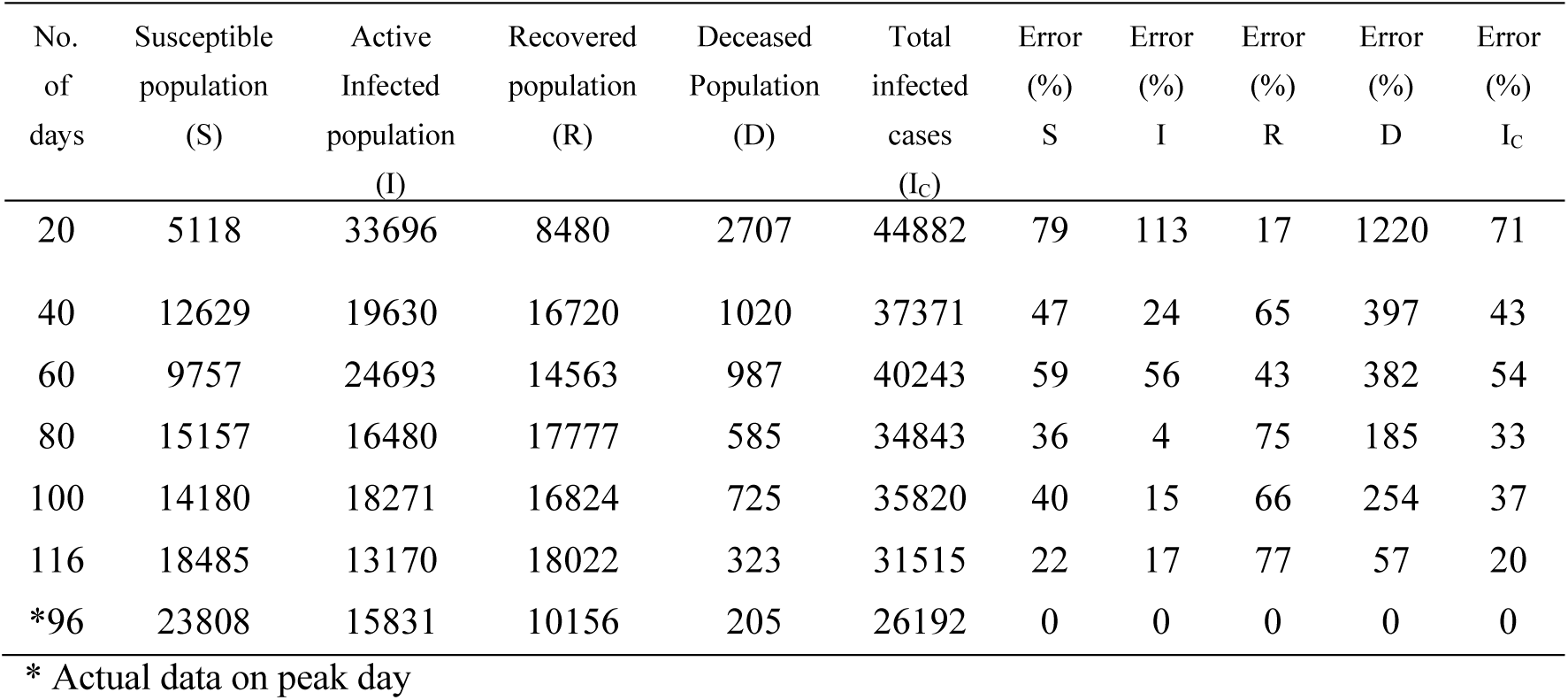
Sensitivity of SIRD results on peak infectious day of COVID-19 in Kuwait (19 June 2020).

| No. of days | Susceptible population (S) | Active Infected population (I) | Recovered population (R) | Deceased Population (D) | Total infected cases (I<sub>c</sub>) | Error (%) S | Error (%) I | Error (%) R | Error (%) D | Error (%) I<sub>c</sub> |
| --- | --- | --- | --- | --- | --- | --- | --- | --- | --- | --- |
| 20 | 5118 | 33696 | 8480 | 2707 | 44882 | 79 | 113 | 17 | 1220 | 71 |
| 40 | 12629 | 19630 | 16720 | 1020 | 37371 | 47 | 24 | 65 | 397 | 43 |
| 60 | 9757 | 24693 | 14563 | 987 | 40243 | 59 | 56 | 43 | 382 | 54 |
| 80 | 15157 | 16480 | 17777 | 585 | 34843 | 36 | 4 | 75 | 185 | 33 |
| 100 | 14180 | 18271 | 16824 | 725 | 35820 | 40 | 15 | 66 | 254 | 37 |
| 116 | 18485 | 13170 | 18022 | 323 | 31515 | 22 | 17 | 77 | 57 | 20 |
| *96 | 23808 | 15831 | 10156 | 205 | 26192 | 0 | 0 | 0 | 0 | 0 |

\* Actual data on peak day

## IV. Conclutions

It is highly important that we estimate accurately COVID-19 dynamics as early as possible. Here, we investigated sensitivity of SIRD model on predicting trends of COVID-19 in Kuwait. We optimized SIRD model parameters so that we get high resolution and best fit to 3-set of COVID-19 data simultaneously for any time duration of data. We used 20, 40, 60, 80, 100, and 116 days data cases separately to fit SIRD model with each time interval. The following conclusion are drawn:

- SIRD model have accurately predicted the peak day of infectious after 40 days development of COVID-19 within 1-10 June 2020 in Kuwait. The actual peak day was 30 May 2020. Sensitivity analysis of SIRD model on peak infectious day indicated that the model is not very accurate with error bands of 22-79% for susceptible, 4-113% for active infected, 17-77% for recovered, 57-1220% for deceased, and 20-71% for total infected.
- SIRD model can be considered as a rough model for predicting number of exposed populations, and better models with more controlling factors may be needed.
- Trends of reducing the reproduction number (R0) since the outbreak of COVID-19 in Kuwait is a promising indicator in slowing down the disease.
- The high value of the reproduction number (R0) is a bad indicator and may lead to the second wave of COVID-19 infection. Results of active infected cases on 26 June 2020 might indicates increasing trends towards the second peak of infectious in Kuwait.

SIRD model is a useful tool for a quick and rough estimation of peak day of infectious for COVID-19 pandemic, yet it is not a precise method. Advanced versions of the SIR model or other new models are needed with more controlling factors to find more precise predictions.

## Data Availability

data is public at Worldometer, https://www.worldometers.info/world-population/kuwaitpopulation/, accessed 26 June 2020.

## References

[1] Worldometer, https://www.worldometers.info/world-population/kuwait-population/, accessed 26 June 2020.

[2] COVID-19 updates, State of Kuwait live, https://corona.e.gov.kw/En/, accessed 26 June 2020.

[3] Sedaghat A, Mostafaeipour N, Abbas Oloomi SA. Prediction of COVID-19 Dynamics in Kuwait using SIRD Model. Integr J Med Sci [Internet]. 2020 Jun. 9 [cited 2020Jun.26];7. Available from: https://www.mbmj.org/index.php/ijms/article/view/170.

[4] Kermack, W. O. and McKendrick, A. G. “A Contribution to the Mathematical Theory of Epidemics.” Proc. Roy. Soc. Lond. A 115, 700–721, 1927.

[5] Devore, Jay L. (2011). Probability and Statistics for Engineering and the Sciences (8th ed.). Boston, MA: Cengage Learning. pp. 508–510. ISBN 978-0-538-73352-6.

[6] Anderson, R. M. and May, R. M. “Population Biology of Infectious Diseases: Part I.” Nature 280, 361–367, 1979.

[7] Weston C. Roda, Marie B. Varughese, Donglin Han, Michael Y. Li, Why is it difficult to accurately predict the COVID-19 epidemic? Infectious Disease Modelling, Volume 5, 2020, Pages 271–281.

[8] Faïçal Ndaïrou, Iván Area, Juan J. Nieto, Delfim F.M. Torres, Mathematical modeling of COVID-19 transmission dynamics with a case study of Wuhan, Chaos, Solitons & Fractals, Volume 135, 2020.

[9] Jones, D. S. and Sleeman, B.D. Ch. 14 in Differential Equations and Mathematical Biology. London: Allen & Unwin, 1983.

[10] Hethcote H (2000). “The Mathematics of Infectious Diseases”. SIAM Review. 42 (4): 599–653. Bibcode:2000, SIAMR.42.599H. doi:10.1137/s0036144500371907.

